# Transmission dynamics and control of COVID-19 in Chile, March-October, 2020

**DOI:** 10.1101/2020.05.15.20103069

**Authors:** Amna Tariq, Eduardo A. Undurraga, Carla Castillo Laborde, Katia Vogt-Geisse, Ruiyan Luo, Richard Rothenberg, Gerardo Chowell

**Affiliations:** Department of Population Health Sciences, School of Public Health, Georgia State University, Atlanta, GA, USA; Escuela de Gobierno, Pontificia Universidad Católica de Chile, Santiago, RM, Chile; Millennium Initiative for Collaborative Research in Bacterial Resistance (MICROB-R), Santiago, RM, Chile; Centro de Epidemiología y Políticas de Salud, Facultad de Medicina, Clínica Alemana Universidad del Desarrollo, Santiago, RM, Chile; Facultad de Ingeniería y Ciencias, Universidad Adolfo Ibáñez, Santiago, RM, Chile

## Abstract

Since the detection of the first case of COVID-19 in Chile on March 3^rd^, 2020, a total of 513188 cases, including ∼14302 deaths have been reported in Chile as of November 2^nd^, 2020. Here, we estimate the reproduction number throughout the epidemic in Chile and study the effectiveness of control interventions especially the effectiveness of lockdowns by conducting short-term forecasts based on the early transmission dynamics of COVID-19. Chile’s incidence curve displays early sub-exponential growth dynamics with the deceleration of growth parameter, *p*, estimated at 0.8 (95% CI: 0.7, 0.8) and the reproduction number, *R*, estimated at 1.8 (95% CI: 1.6, 1.9). Our findings indicate that the control measures at the start of the epidemic significantly slowed down the spread of the virus. However, the relaxation of restrictions and spread of the virus in low-income neighborhoods in May led to a new surge of infections, followed by the reimposition of lockdowns in Greater Santiago and other municipalities. These measures have decelerated the virus spread with *R* estimated at ∼0.96(95% CI: 0.95, 0.98) as of November 2^nd^, 2020. The early sub-exponential growth trend (*p* ∼0.8) of the COVID-19 epidemic transformed into a linear growth trend (*p* ∼0.5) as of July 7^th^, 2020, after the reimposition of lockdowns. While the broad scale social distancing interventions have slowed the virus spread, the number of new COVID-19 cases continue to accrue, underscoring the need for persistent social distancing and active case detection and isolation efforts to maintain the epidemic under control.

**Author summary:** In context of the ongoing COVID-19 pandemic, Chile has been one of the hardest-hit countries in Latin America, struggling to contain the spread of the virus. In this manuscript, we employ renewal equation to estimate the reproduction number (*R*) for the early ascending phase of the COVID-19 epidemic and by July 7^th^, 2020 to guide the magnitude and intensity of interventions required to combat the COVID-19 epidemic. We also estimate the instantaneous reproduction number throughout the epidemic in Chile. Moreover, we generate short-term forecasts based on the epidemic trajectory using phenomenological models, and assess counterfactual scenarios to understand any additional resources required to contain the virus’ spread. Our results indicate early sustained transmission of SARS-CoV-2. However, the initial control measures at the start of the epidemic significantly slowed down the spread of the virus. The easing of COVID-19 restrictions in April led to a new wave of infections, followed by the re-imposition of lockdowns in Greater Santiago and other municipalities. Most recent estimates of reproduction number indicate a decline in the virus transmission. While broad-scale social distancing interventions have slowed the virus spread, the number of new COVID-19 cases continue to accrue, underscoring the need for persistent social distancing efforts.

## Introduction

The coronavirus disease 2019 (COVID-19), caused by severe acute respiratory syndrome coronavirus 2 (SARS-CoV-2), was declared a global pandemic by the World Health Organization (WHO) on March 11^th^, 2020 [1, 2]. This highly contagious unprecedented virus has impacted government and public institutions, strained the health care systems, restricted people in their homes, and caused country-wide lockdowns resulting in a global economic crisis [3-5]. Moreover, as of November 2^nd^, 2020, nearly 46 million COVID-19 cases in 213 countries and territories have been reported, including more than 1.2 million deaths [6]. The social, economic, and psychological impact of this pandemic on much of the world’s population is profound [7-13].

Soon after its initial rapid spread in China, the first case of novel coronavirus beyond China was reported in Thailand on January 13^th^, 2020 [14]. The first case in the USA was not identified until January 20^th^, 2020 followed by the detection of the first cases in the European territory on January 24^th^, 2020 [15, 16]. The COVID-19 pandemic has since spread to every continent except the Antarctica. While some countries like New Zealand and Australia have steadily suppressed the COVID-19 spread, reporting less than 150 cases per day as of November 2^nd^, 2020, other countries like Brazil, India, and the USA still struggle to contain the increasing number of cases [17]. Subsequently, considerable COVID-19 outbreaks have occurred in Latin America since late February 2020.

The WHO declared Latin America the new epicenter of the COVID-19 on May 22^nd^, 2020 [18]. Latin America has paid a high toll during the COVID-19 pandemic, with some of the worlds’ highest death rates [19-21]. While home to less than 10% of the world population, Latin America accounts for about one-third of all reported global deaths (∼370 thousand) [6]. Several socioeconomic, demographic, and political factors make control of the pandemic in Latin America particularly challenging [22-25]. Most countries in the region are now facing the stark social and economic costs imposed by large-scale non-pharmaceutical interventions while largely failing to control the epidemic’s spread [13, 24, 26]. Despite these unique conditions, the region has received relatively little attention from researchers globally [19]. As of November 1^st^, 2020, the highest number of cases have been reported in Brazil (5,516,658), followed by Argentina (1,157,179), Colombia (1,063,151), Mexico (918,811), Peru (900,180) and Chile (510,256) [17, 27]. Adjusted by population, Chile’s COVID-19 outbreak is among the worst globally, with more than 26,000 cases and 980 deaths per million inhabitants [28].

The first case of SARS-CoV-2 in Chile was identified on March 3^rd^, 2020. While the initial cases were imported from southeast Asia and Europe, the COVID-19 case counts have expanded in this country, placing Chile in phase 4 of the pandemic on March 25^th^, 2020 [28, 29]. Chile was the fifth country in Latin America after Brazil, Mexico, Ecuador and Argentina to report COVID-19 cases. The first six imported cases were reported in Talca and in the capital of Chile, Santiago [28]. However, since the early phase of the outbreak, Chile has employed an agile public health response by announcing a ban on public health gatherings of more than 500 people on March 13^th^, 2020, when the nationwide cumulative case count reached 44 reported cases [30].

Moreover, the Chilean government announced the closure of all daycares, schools, and universities on March 16^th^, 2020. These closures were followed by the announcement to close country borders on March 18^th^, 2020, and the declaration of national emergency on the same date, accompanied by several concrete interventions to further contain the outbreak in the region [31]. In particular, these included a night-time curfew in Chile starting on March 22^nd^, 2020, and localized lockdowns (i.e., intermittent lockdowns at the municipality level depending on total cases and case growth) starting on March 28^th^, 2020 in two municipalities in Southern Chile and seven municipalities in Santiago [32]. These initial containment strategies kept the COVID-19 case counts lower than regional peers; Brazil, Peru, and Ecuador until the end of April 2020. However, the government started to ease the COVID-19 restrictions in late April by reopening the economy under the “Safe Return” plan, including the televised opening of some businesses and stores, as new infections had reduced between 350-500 per day by the end of April, implying an only apparent flattening of the COVID-19 curve [33-35]. Moreover, imposing and lifting lockdowns in small geographical areas (municipalities) proved unsuccessful in areas with high interdependencies such as the Greater Santiago [36]. This strategy resulted in a new wave of infections; with the virus spreading from more affluent areas of Chile to more impoverished, crowded communities, forcing the government to reimpose lockdown measures in Santiago in mid-May (Figure 1) [23, 37, 38]. By mid-July, the government implemented the “step by step” strategy, considering five stages of gradual opening, at the municipality level, based on the periodic monitoring of epidemiological and health system indicators. The case counts continued to increase, averaging ∼4943 cases per day in June 2020, and started to decline thereafter. The mid-June peak of infections resulted in intensive care units (ICU) reaching saturation levels of 89% nationally and 95% in the Metropolitan Region [39]. Thus far, Chile has accumulated 513,188 reported cases including 14,302 deaths as of November 2^nd^, 2020. The majority (∼52%) of COVID-19 cases are concentrated in Region Metropolitana (mostly in Chile’s capital, Santiago), with 297,423 reported cases, followed by 30,498 cases in Valparaiso located in coastal central Chile, and 30,934 cases in Biobio located in southern Chile [40, 41]. Moreover, the crude case fatality rate in Chile (∼2.8%) resonates with the global average case fatality rate (2.6%) [17, 42].

**Figure 1:**
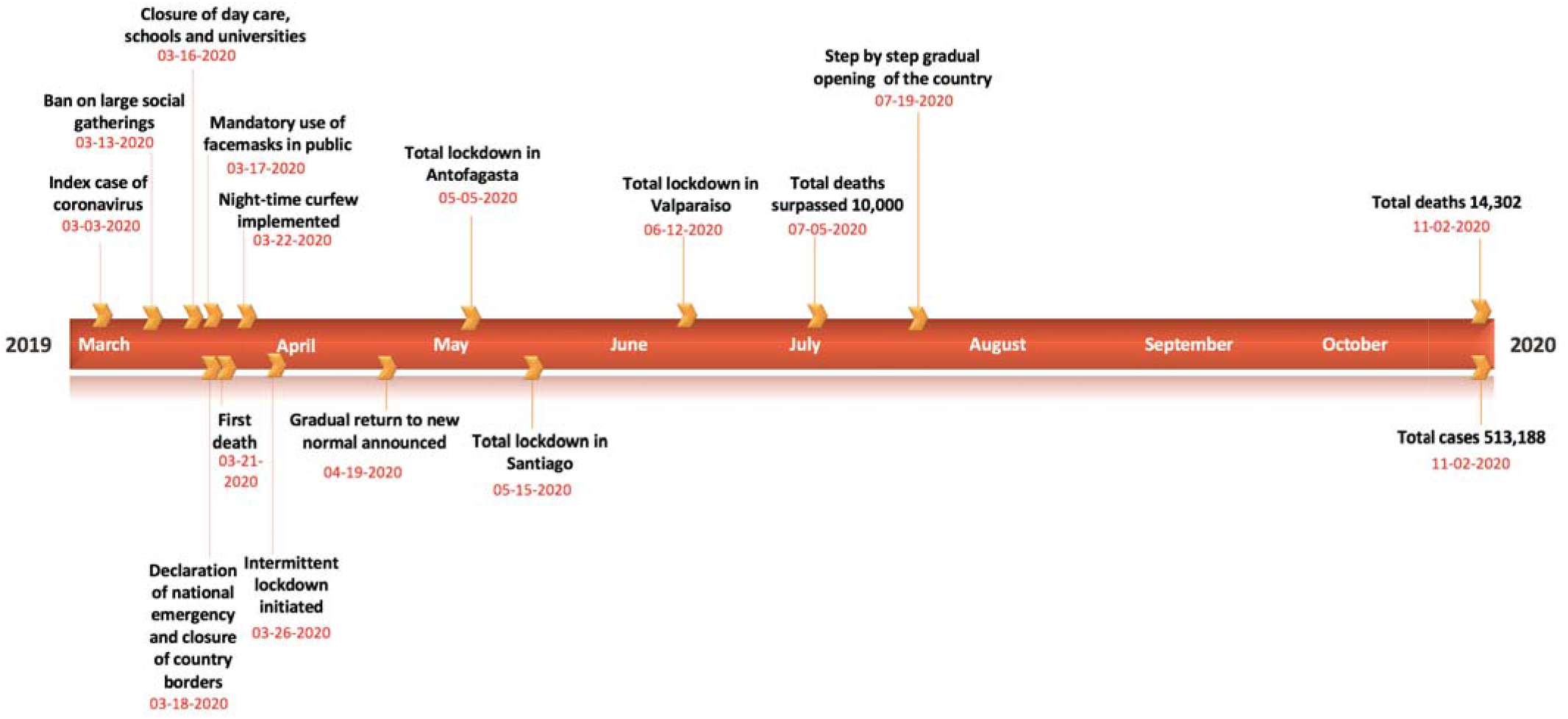
Timeline of the milestones COVID-19 pandemic in Chile as of November 2^nd^, 2020.

In this study, we estimate the transmission potential of COVID-19, including the effective reproduction number, *R*, during the early transmission phase of the COVID-19 epidemic in Chile and around the mid of the epidemic, by July 7^th^, 2020. We also estimate the instantaneous reproduction number throughout the epidemic in Chile. The reproduction number can guide the magnitude and intensity of control interventions required to combat the COVID-19 outbreak [43, 44]. We examine the effectiveness of control interventions in Chile (see Table 1) on the transmission rate. To do this, we conduct short-term forecasts using phenomenological growth models calibrated using the early trajectory of the epidemic and by the mid of the epidemic (as of July 7^th^, 2020) [45] to anticipate additional resources required to contain the epidemic. These phenomenological growth models are useful in capturing the epidemic’s empirical patterns, especially when the epidemiological data are limited, and significant uncertainty exists around infectious disease epidemiology [46]. These models provide a starting point for forecasting the epidemic size and characterizing the temporal changes in the reproduction number during the epidemic [47].

**Table 1:** Timeline of the implementation of the social distancing interventions in Chile as of November 2^nd^, 2020.

| Date | Control interventions |
| --- | --- |
| March 13<sup>th</sup>, 2020 | Ban on large social gatherings implemented in Chile [30] |
| March 16<sup>th</sup>, 2020 | Closures of daycares, schools, and universities in Chile [32, 48]<br>Mandatory quarantine of high-risk individuals returning from Iran, China, West Europe and South Korea |
| March 18<sup>th</sup>, 2020 | Declaration of national emergency (14)<br>Closure of country borders (14)<br>Telework implemented |
| March 19<sup>th</sup>, 2020 | Closure of mall and department stores with the exception of supermarkets, pharmacies, banks and grocery stores |
| March 21<sup>st</sup>, 2020 | Closure of non-essential business including theatres, restaurant, bars and gyms |
| March 22<sup>nd</sup>, 2020 | Night time curfew implemented [32] |
| March 26<sup>th</sup>, 2020 | Intermittent lockdown initiated (implemented at municipality level) [32] |
| April 8<sup>th</sup>, 2020 | Orders on mandatory use of facemasks in public transport [49] |
| April 17<sup>th</sup>, 2020 | Orders on mandatory use of facemasks in all public spaces [60] |
| April 30<sup>th</sup>, 2020 | First shopping mall is reopened in Santiago and then closed the next day [50] |
| May 5<sup>th</sup>, 2020 | Total lockdown in Antofagasta [31] |
| May 15<sup>th</sup>, 2020 | Total lockdown imposed in all municipalities of Santiago [51] |
| June 12<sup>th</sup>, 2020 | Total lockdown in Valparaiso [31] |
| July 19<sup>th</sup>, 2020 | Step by step gradual reopening of the country |

## Methods

### COVID-19 incidence and testing data

We obtained updates on the daily series of new COVID-19 cases as of November 2^nd^, 2020, from the publicly available data from the GitHub repository created by the Chile’s government [27]. Incidence case data by the date of reporting per day, confirmed by PCR (polymerase chain reaction) tests from March 3^rd^–November 2^nd^, 2020, were analyzed. The daily testing and positivity rates available from April 9^th^–November 2^nd^, 2020, were also analyzed.

### Models

We utilize two phenomenological growth models, the generalized growth model (GGM) and the generalized logistic growth model (GLM) that have been validated by deriving short-term forecasts for multiple infectious diseases in the past, including SARS, pandemic Influenza, Ebola, and Dengue [52, 53].

### Generalized growth model (GGM)

We generate short term forecasts using the generalized growth model (GGM) that characterizes the early ascending phase of the epidemic by estimating two parameters: (1) the intrinsic growth rate, *r*; and (2) a dimensionless “deceleration of growth” parameter, *p*. This model allows to capture a range of epidemic growth profiles by modulating parameter *p*. The GGM model is given by the following differential equation:

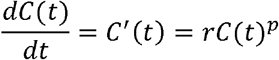

In this equation *C′(t)* describes the incidence curve over time *t, C(t)* describes the cumulative number of cases at time *t* and *p*∈[0,1] is a “deceleration of growth” parameter. This equation becomes constant incidence over time if *p*=0 and an exponential growth model for cumulative cases if *p* =1. Whereas if *p* is in the range 0< *p* <1, then the model indicates sub-exponential growth dynamics [54, 55].

### Generalized logistic growth model (GLM)

The generalized logistic growth model (GLM) is an extension of the simple logistic growth model that captures a range of epidemic growth profiles, including sub-exponential (polynomial) and exponential growth dynamics. GLM characterizes epidemic growth by estimating (i) the intrinsic growth rate, *r* (ii) a dimensionless “deceleration of growth” parameter, *p* and (iii) the final epidemic size, *k*_0_. The final epidemic size is sensitive to small variations in the deceleration of growth parameters [56] and would vary as the epidemic progresses. The deceleration parameter modulates the epidemic growth patterns, including the sub-exponential growth (0< *p* <1), constant incidence (*p* =0) and exponential growth dynamics (*p* =1). The GLM model is given by the following differential equation:

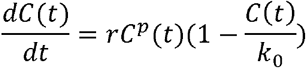

Where 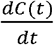 describes the incidence over time *t* and the cumulative number of cases at time *t* is given by *C(t)* [45]. This simple logistic growth type model typically supports single peak epidemics in the number of new infections followed by a burnt-out period, unless external driving forces such as the seasonal variations in contact patterns exist. This model can underestimate the peak timing and the duration of outbreaks. This model can also underestimate the case incidence before the inflection point has occurred [45, 47, 53, 57].

### Calibration of the GGM and GLM model

We calibrate the GGM and the GLM model to the daily incidence curve by dates of reporting in Chile using time series data from March 3^rd^–March 30^th^, 2020, and from May 9^th^ – July 7^th^, 2020, respectively (Figure 2). The period from March 3^rd^–March 30^th^, 2020, includes the initial interventions made by the Chilean government, whereas the period from May 9^th^-July 7^th^, 2020, comprises the reimposition of lockdowns after a brief reopening of society under the “new normal” (Figure 1).

**Figure 2:**
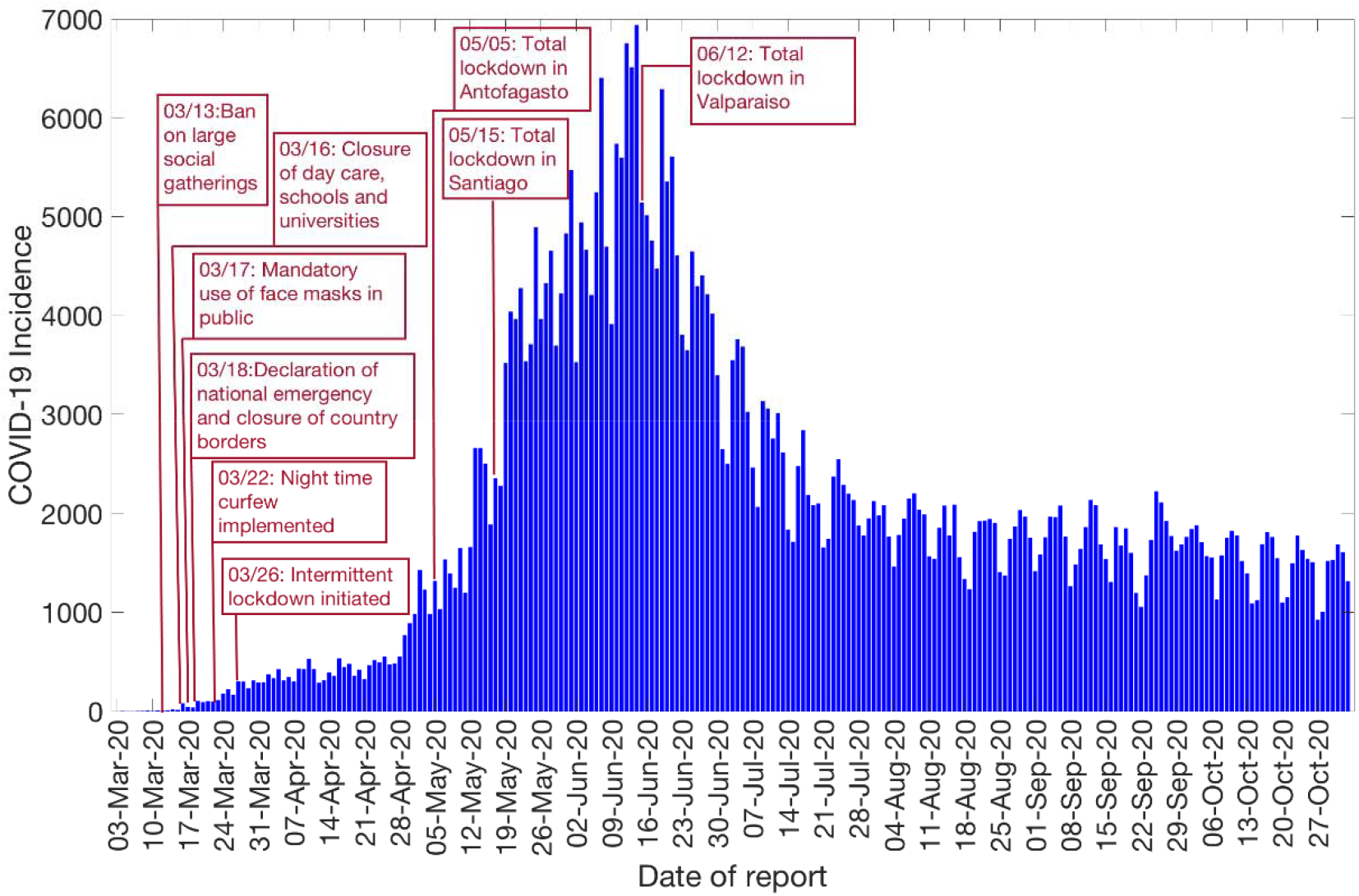
Daily incidence curve for all COVID-19 confirmed cases in Chile as of November 2^nd^, 2020 (9).

Model parameters are estimated by a non-linear least-square fitting of the model solution to the incidence data by the date of reporting. This is achieved by searching for the set of model parameters 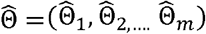 that minimizes the sum of squared differences between the observed data *y*_*ti*_ = *y*_*t1*_, *y*_*t2*_,*…. y*_*tn*_ and the corresponding mean incidence curve given by 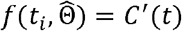 where 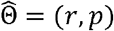 corresponds to the set of parameters of the GGM model and 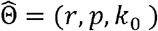 corresponds to the set of parameters of the GLM model. In both cases, the objective function for the best fit solution of 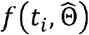 is given by :

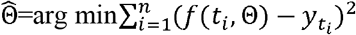

where *t*_*i*_ is the time stamp at which the time series data are observed and n is the total number of data points available for inference. The initial condition is fixed to the first observation in the data set. This way, 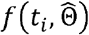 gives the best fit to the time-series data 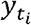 Next, we utilize a parametric bootstrapping approach assuming a negative binomial error structure for the GGM and GLM model to derive uncertainty in the parameters obtained by non-linear least-square fit of the data as previously described [54, 58]. The variance is assumed to be three times the mean for GGM and 96 times the mean for the GLM. The model confidence intervals of parameters and the 95% prediction intervals of model fit are also obtained using the parametric bootstrap approach [54].

### Reproduction number, *R*, from case incidence using GGM

The reproduction number, *R*, is defined as the average number of secondary cases generated by a primary case at time *t* during the outbreak. This is crucial to identify the intensity of interventions required to contain an epidemic [59-61]. Estimates of effective *R* indicate if the disease transmission continues (*R*>1) or if the active disease transmission ceases (*R*<1). Therefore, in order to contain an outbreak, we need to maintain *R*<1. We estimate the reproduction number by calibrating the GGM to the epidemic’s early growth phase (27 days) [55]. We model the generation interval of SARS-CoV-2, assuming gamma distribution with a mean of 5.2 days and a standard deviation of 1.72 days [62]. We estimate the growth rate parameter, *r*, and the deceleration of growth parameter, *p*, as described above. The progression of local incidence cases I_i_ at calendar time t_i_ is simulated from the calibrated GGM model. Then in order to estimate the reproduction number, we apply the discretized probability distribution of the generation interval denoted by *ρ*_*i*_ to the renewal equation as follows [43, 44, 63]:

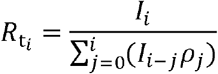

The numerator represents the total new cases *I*_*i*_, and the denominator represents the total number of cases that contribute to generating the new cases *I*_*i*_ at time *t*_*i*_. Hence, *R*_*t*_, represents the average number of secondary cases generated by a single case at time *t*. Next, we derive the uncertainty bounds around the curve of *R*_*t*_ directly from the uncertainty associated with the parameter estimates (*r, p*) obtained from the GGM. We estimate *R*_*t*_ for 300 simulated curves assuming a negative binomial error structure where the variance is assumed to be three times the mean [54].

### Reproduction number, *R*, from case incidence using GLM

In order to estimate the reproduction number by July 7^th^, 2020 (after the reimposition of lockdowns in Santiago and Valparaiso), we calibrate the GLM from May 9^th^ – July 7^th^, 2020 [55]. Next, we model the generation interval [62], estimate the model parameters (*r, p, k*_0_) from GLM and the reproduction number from the renewal equation as described above [43, 44, 63]. The uncertainty bounds around the curve of *R*_*t*_ are derived directly from the uncertainty associated with the parameter estimates (*r, p, k*_0_). We estimate *R*_*t*_ for 300 simulated curves assuming a negative binomial error structure [54] where the variance is assumed to be 96 times of the mean calculated by averaging mean to variance ratio calculated from the data (by binning data points and calculating directly from the data itself).

### Instantaneous reproduction number, *R*, using the Cori method

We estimate *R* by the ratio of number of new infections generated at time *t* (*I*_*t*_), to the total infectiousness of infected individuals at time t, given by :

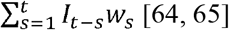

In this equation, *w*_*s*_ represents the infectivity profile of the infected individual, which depends on the time since infection (*s)*, but is independent of the calendar time (*t*) [66, 67]. More specifically, *w*_*s*_ is defined as a probability distribution describing the average infectiousness profile of an individual after infection. Distribution of *w*_*s*_ is affected by individual biological factors such as symptom severity or pathogen shedding. The equation 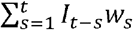 indicates the sum of infection incidence up to time step *t* − 1, weighted by the infectivity function *w*_*s*_. The distribution of the generation time can be utilized to approximate the infectivity profile, *w*_*s*_, however, since the time of infection is rarely observed, it becomes difficult to measure the distribution generation time [64]. Hence, time of symptom onset is usually used to estimate the distribution of serial interval, which is defined as the time interval between the dates of symptom onset among two successive cases in a transmission chain [68]. The infectiousness of a case is a function of the time since infection. This quantity is proportional to *w*_*s*_ if we set the timing of infection in the primary case as the time zero of *w*_*s*_ and assume that the generation interval equals the SI. The SI was assumed to follow a gamma distribution with a mean of 5.2 days and a standard deviation of 1.72 days [62]. Analytical estimates of *R*_*t*_ were obtained within a Bayesian framework using EpiEstim R package in R language [68]. *R*_*t*_ was estimated at 7-day intervals. We reported the median and 95% credible interval (CrI).

## 3. Results

### Case incidence data

The Ministry of Health Chile reported a total of 481,342 COVID-19 cases as of November 2^nd^, 2020 [27]. The epidemic curve showed an increasing trajectory from April-June 2020 and declined thereafter. On average, ∼443 (SD: 133.6) new cases per day were reported in April 2020, ∼2697 (SD:1342) new cases per day were reported in May 2020 and ∼4943 (SD:972.2) new cases per day were reported in June 2020, the maximum number of cases reported per day during the epidemic. The per-day cases declined starting July, with ∼2456 (SD:581) new cases reported per day in July 2020, ∼1808 (SD:258) new cases per day reported in August 2020, ∼1706 (SD:294) new cases per day reported in September 2020, and ∼1521 (SD:275) new cases per day reported in October 2020. Figure 2 shows the daily incidence data of all confirmed cases in Chile as of November 2^nd^, 2020.

### Initial growth dynamics and estimate of the reproduction number using GGM

We estimate the reproduction number for the first 27 epidemic days incorporating the effects of the social distancing interventions, as explained in Table 1 and Figure 1. The incidence curve displays sub-exponential growth dynamics with the scaling of growth parameter (deceleration of growth parameter), *p*, estimated at 0.77 (95% CI: 0.73, 0.81) and the intrinsic growth rate, *r*, estimated at 0.81 (95% CI: 0.67, 1.0). During the early transmission phase the reproduction number was estimated at 1.8 (95% CI: 1.6, 1.9) (Figure 3).

**Figure 3:**
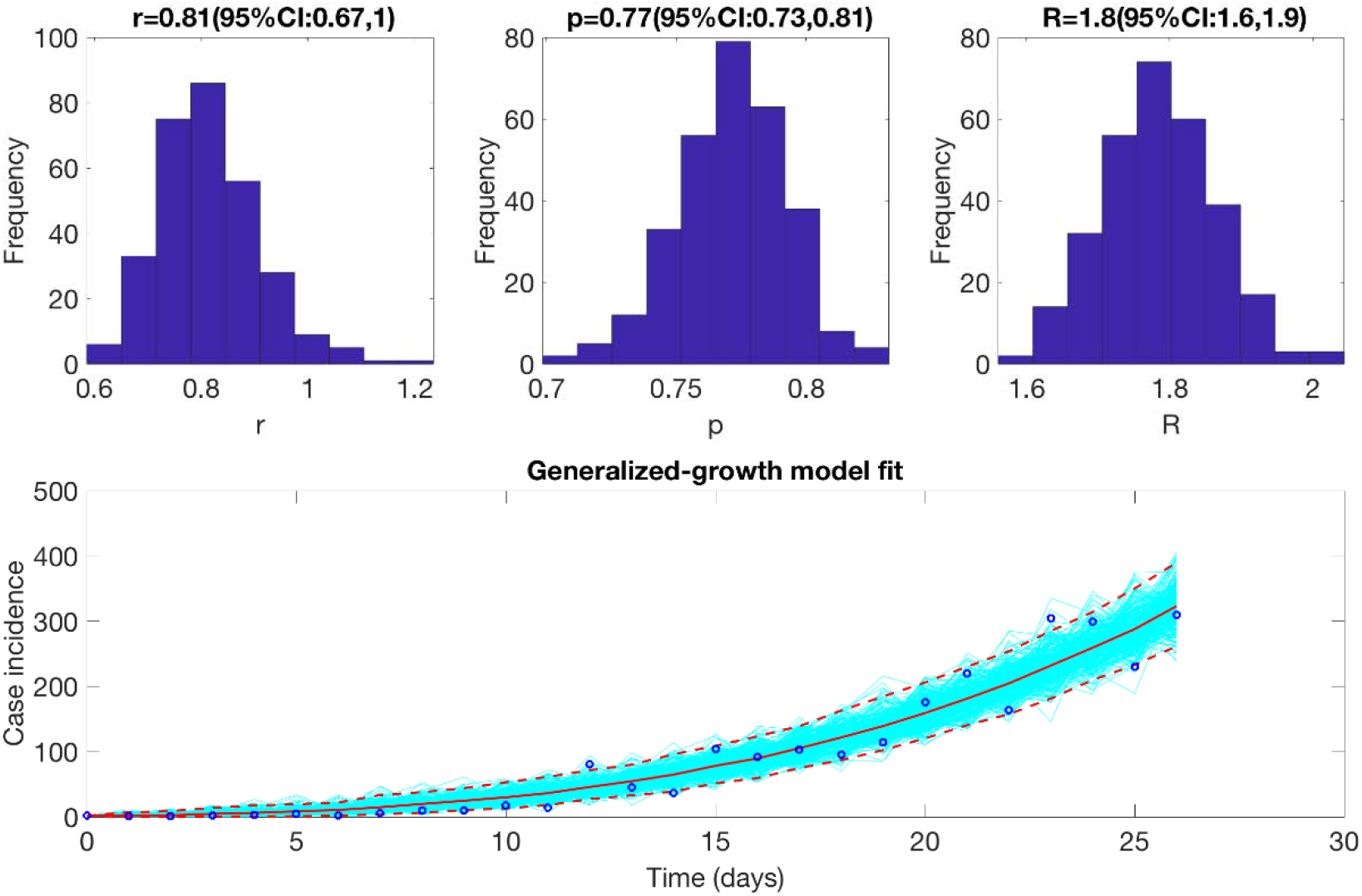
Reproduction number with 95% CI estimated using the GGM model. The estimated reproduction number of the COVID-19 epidemic in Chile as of March 28^th^, 2020, is 1.8 (95% CI: 1.6, 1.9).

### Growth dynamics and estimate of reproduction number using GLM by July 7, 2020

We also estimate the reproduction number from May 9^th^-July 7^th^, 2020, incorporating the effects of the reimplementation of localized lockdowns in Santiago, Antofagasta, and Valparaíso. The incidence curve displays a nearly linear growth trend with the deceleration of growth parameter, *p*, estimated at 0.51 (95% CI: 0.47, 0.56). The deceleration parameter in the GLM model helps modulate the trajectory of the epidemic, depicting a linear growth trend. The intrinsic growth rate, *r*, was estimated at 22 (95% CI: 13, 31) and the final epidemic size, *k*_0_, estimated at 3.4 e+05 (95% CI: 3.1 e+05, 3.7 e+05). The reproduction number was estimated at 0.87 (95% CI: 0.84, 0.89) as of July 7^th^, 2020 (Figure 4).

**Figure 4:**
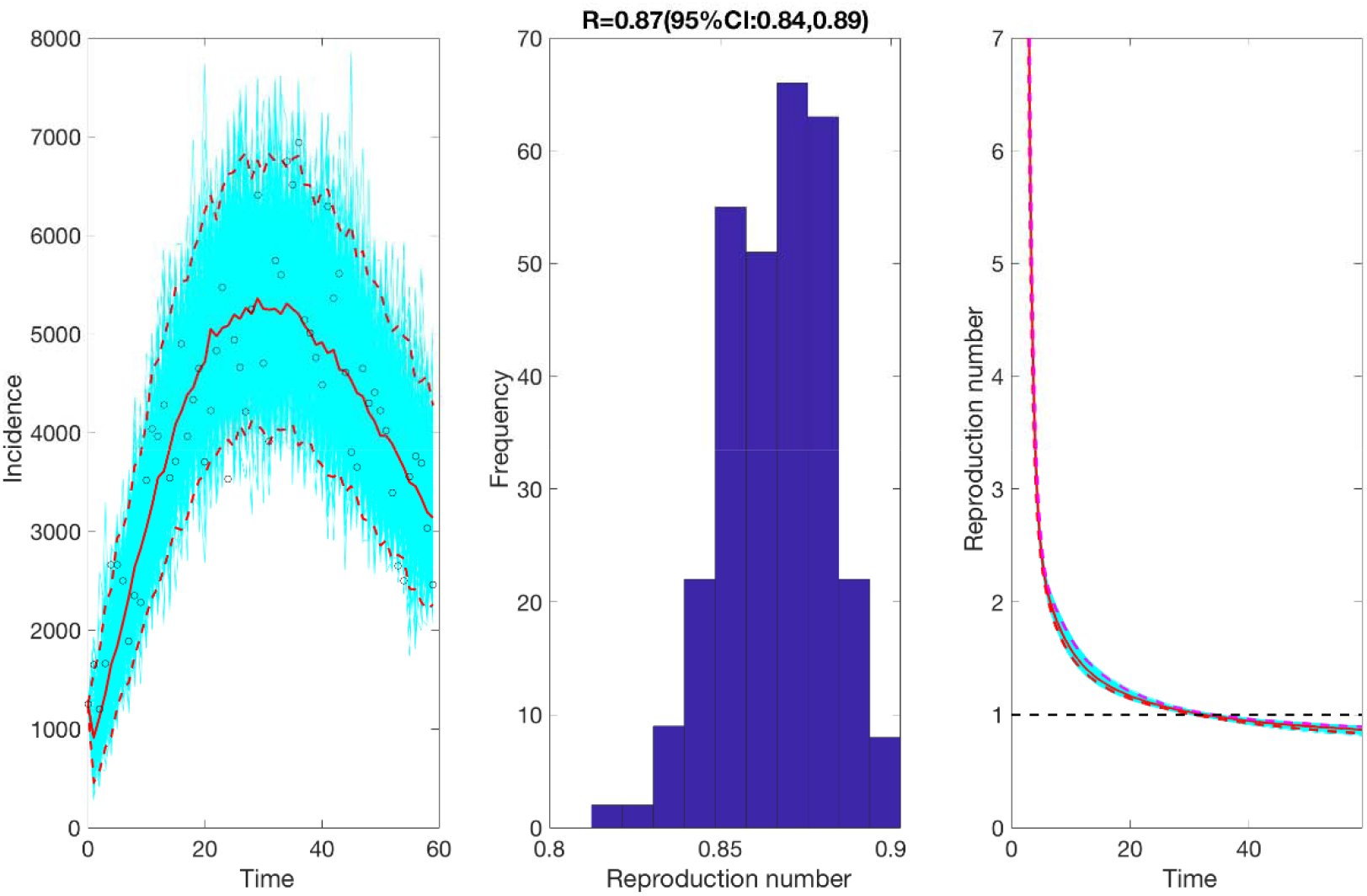
Reproduction number with 95% CI estimated by calibrating the GLM model from May 9^th^-July 7^th^, 2020. The estimated reproduction number of the COVID-19 epidemic in Chile as of July 7^th^, 2020, is 0.87 (95% CI: 0.84, 0.89).

### Estimate of instantaneous reproduction number using Cori method

Utilizing the Cori method based on a sliding weekly window, we observe that the reproduction number peaked on March 16^th^, 2020, with an estimate of R∼ 6.19 (95% CrI= 5.84, 7.08). The reproduction number declined thereafter and reached ∼1.00 (95% CrI: 0.99, 1.04) on April 17^th^, 2020. From April 18^th^-June 18^th^, 2020 the reproduction number fluctuated between 1.01-1.75. This was followed by a decline in the reproduction number to less than 1.0 between June 19^th^-August 9^th^, 2020. Since then, the reproduction number has fluctuated around 1.0 with the most recent estimate of *R* ∼ 0.96 (95% CrI: 0.95, 0.98) (Figure 5).

**Figure 5:**
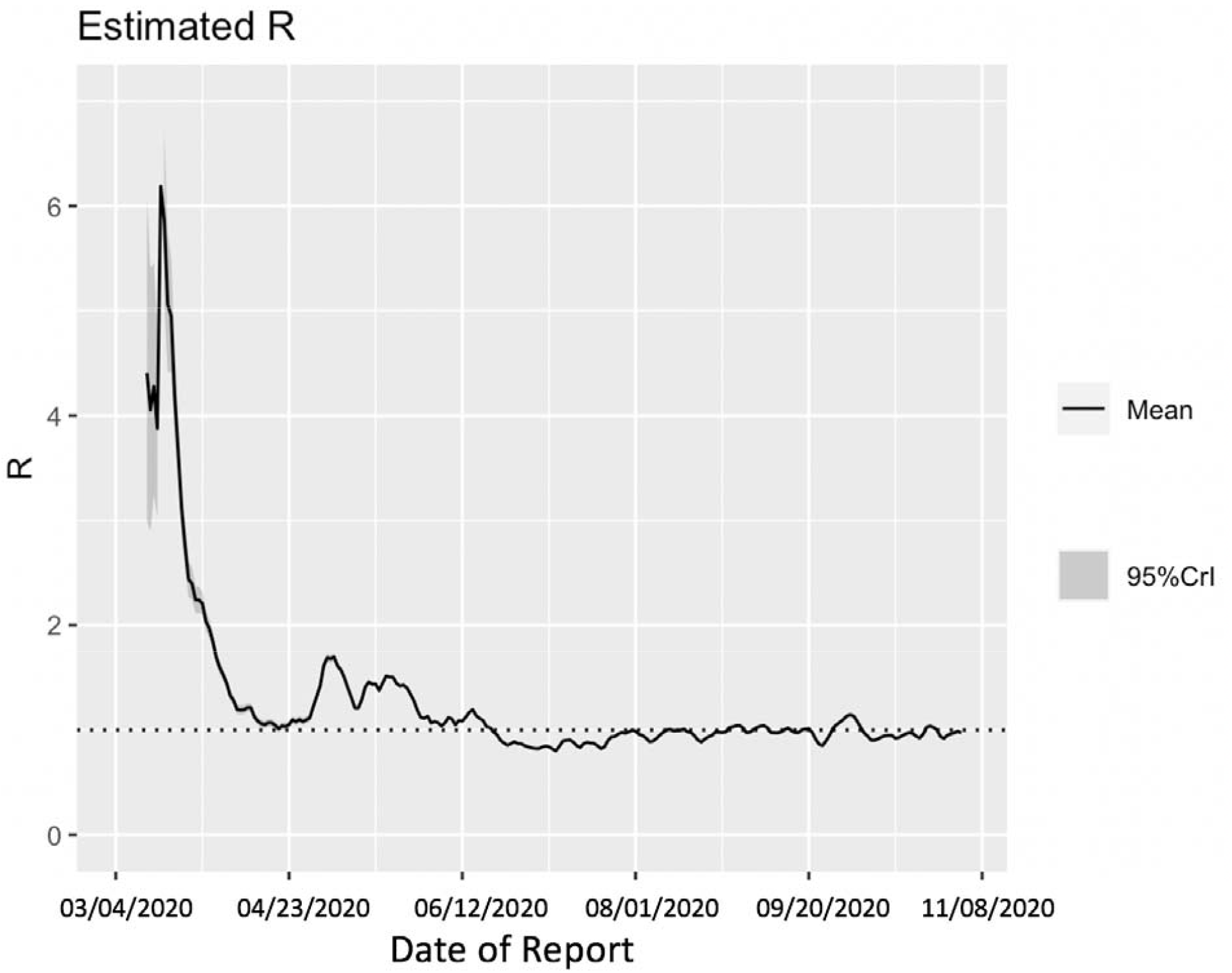
Estimate of instantaneous reproduction number (*R*) for the COVID-19 epidemic in Chile as of November 2^nd^, 2020 using the Cori method. The most recent estimate of *R*∼ 0.96 (95% CrI: 0.95, 0.98) a of November 2^nd^, 2020. Black solid line represents the mean *R* and the gray shaded region represents th 95% credible interval around mean *R*.

### Assessing the impact of social distancing interventions

To assess the impact of social distancing interventions in Chile given in Table 1, we generated a 20-day ahead forecast for Chile based on the daily incidence curve until March 30^th^, 2020. The 28-day calibration period of the GGM model yields an estimated growth rate, *r*, at 0.8 (95% CI: 0.6, 1.0) and a deceleration of growth parameter, *p*, at 0.8 (95% CI: 0.7, 0.8), indicating early sub-exponential growth dynamics. The 20-day ahead forecast suggested that the early social distancing measures significantly slowed down the early spread of the virus in Chile, whose effect is noticeable about two weeks after implementing an intervention, as shown in Figure 6. A case resurgence was observed in Chile in mid-May 2020. As a consequence of this case resurgence, a total lockdown was imposed in Greater Santiago (representing ∼52% of total COVID-19 cases during the epidemic) on May 15^th^, 2020. The quarantine in Santiago was gradually eased from August 17, 2020, and was lifted on September 28, 2020, as a part of the move to phase three of a five-step plan of deconfinement that would allow movement on regional transportation and reopening of non-essential businesses and schools [31, 69, 70]. We generated a 20-day ahead forecast based on the daily incidence curve from May 9^th^-July 7^th^, 2020. The 60-day calibration of the GLM model yields an estimated scaling of the growth parameter, *p*, at 0.52 (95% CI: 0.47, 0.57), representing an almost linear growth pattern. The 20-day ahead average forecast utilizing the GLM model showed that Chile could accumulate ∼45,160 cases (95% CI: 27,934-67,600) between July 8^th^-July 27^th^, 2020 (Figure 7). Our forecast results approximate closely the ∼46798 cases reported between July 8^th^-July 27^th^, 2020 by the Ministry of Health, Chile.

**Figure 6:**
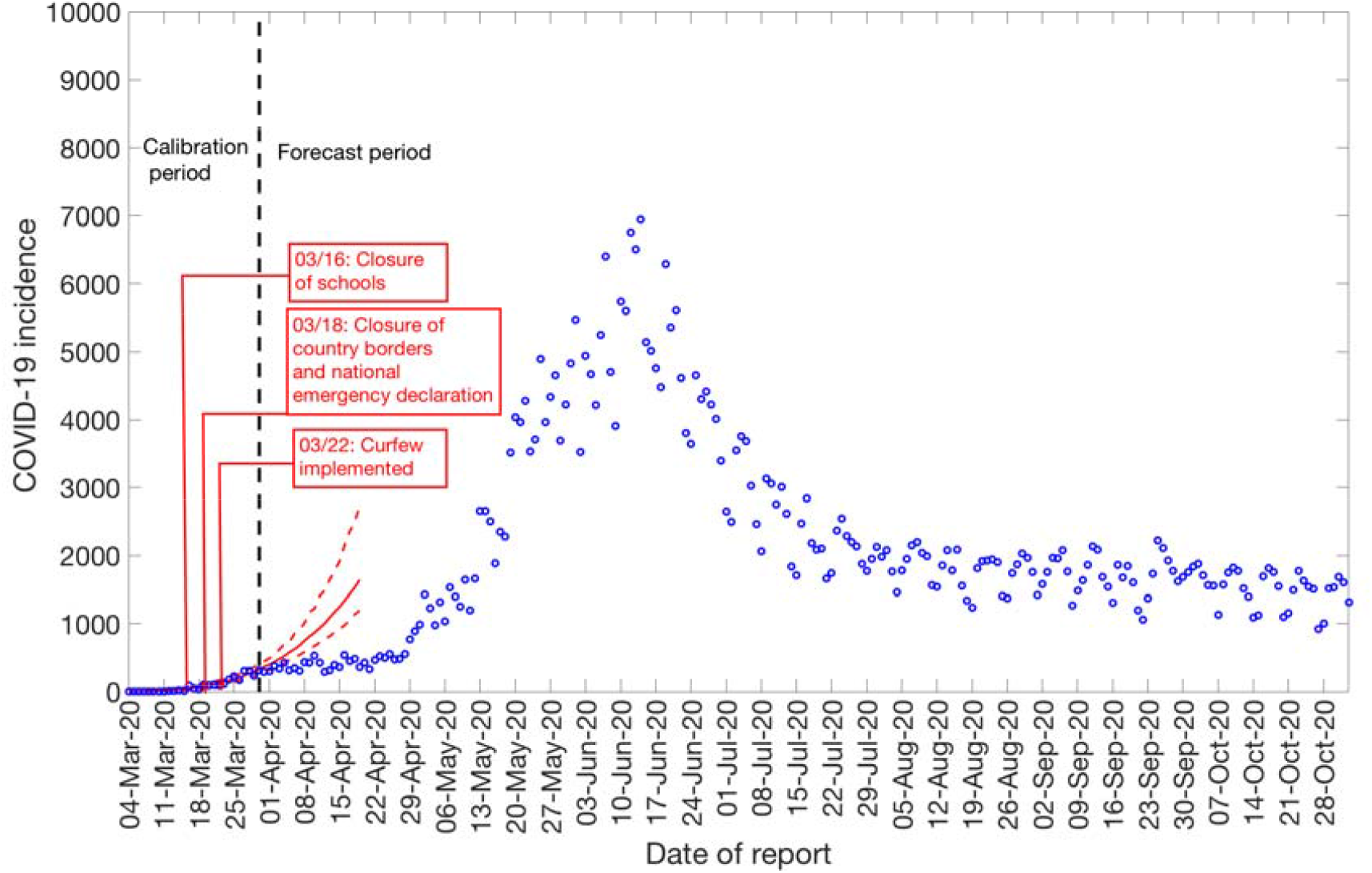
20-days ahead forecast of the COVID-19 epidemic in Chile by calibrating the GGM model until March 30^th^, 2020. Blue circles correspond to the data points; the solid red line indicates the best model fit, and the red dashed lines represent the 95% prediction interval. The vertical black dashed line represents the time of the start of the forecast period.

**Figure 7:**
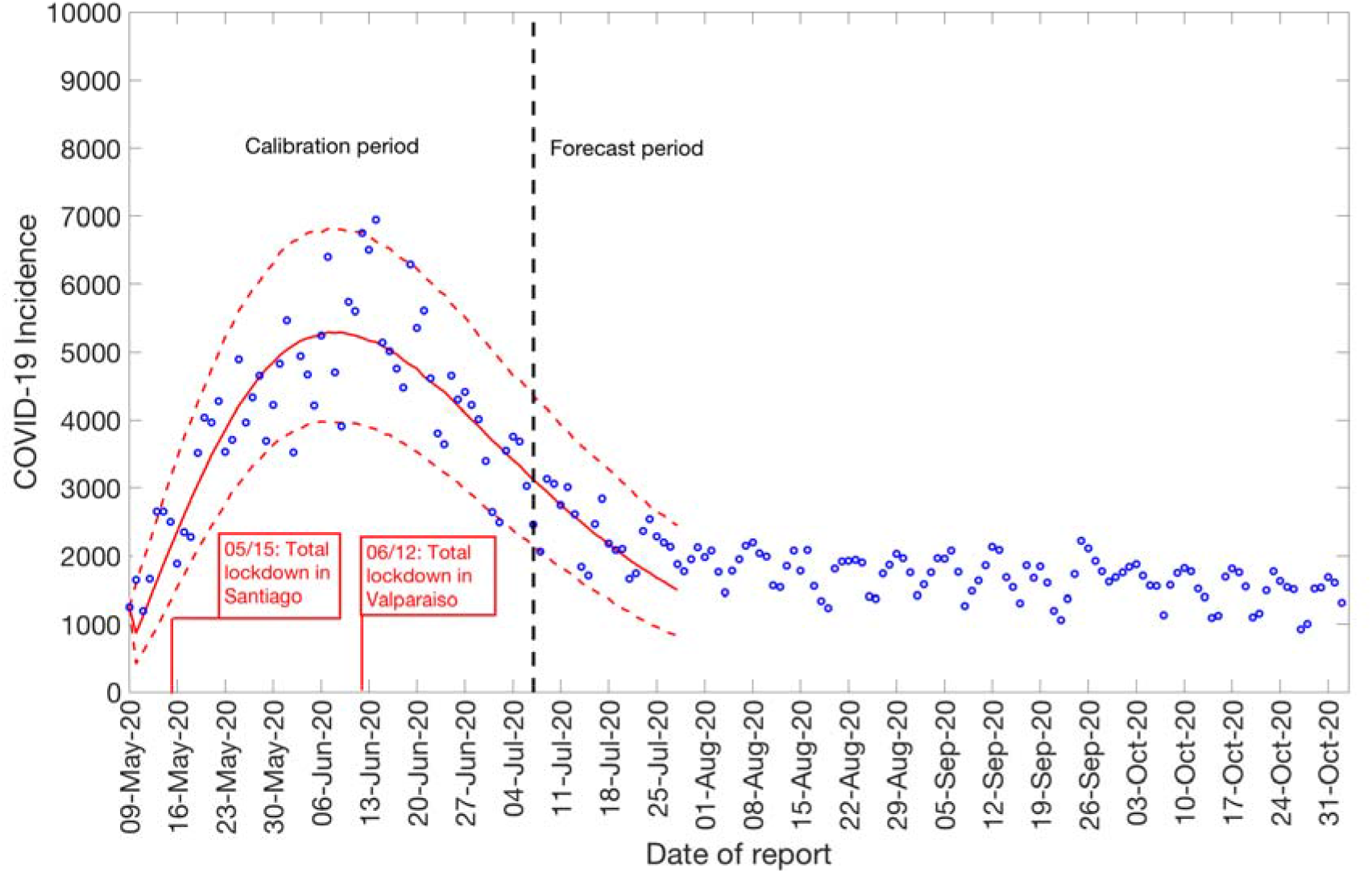
20-days ahead forecast of the COVID-19 epidemic in Chile by calibrating the GLM model from May 9^th^-July 7^th^, 2020. Blue circles correspond to the data points; the solid red line indicates the best model fit, and the red dashed lines represent the 95% prediction interval. The vertical black dashed line represents the time of the start of the forecast period. (96.2 is variance)

### COVID-19 Testing rates and positivity rate

Daily testing and positivity rates for the time period April 9^th^–November 2^nd^, 2020, by the reporting date are shown in Figure 8. The total number of tests performed for this time period were 4,325,617, amongst which 10.9% (47,597) had positive results. The average number of daily tests was estimated at ∼5,460 for April 2020 and ∼12,959 for May 2020, a 137% increase. The testing rate in Chile further increased in June 2020, testing on average ∼17,578 individuals per day, followed by a slight decline in July 2020, testing on average 16587 individuals per day. However, the testing rates continued to increase in August (average ∼26,079 tests per day), September (average ∼29,663 tests per day), and October (average ∼31,821 tests per day), indicating an expanding testing capacity of the country. The positivity rate (percentage of positive tests among the total number of tests) has fluctuated from a monthly average of ∼9.07% (SD: 2.3) in April 2020 to a monthly average of ∼4.87% (SD: 0.65) in October 2020.

**Figure 8:**
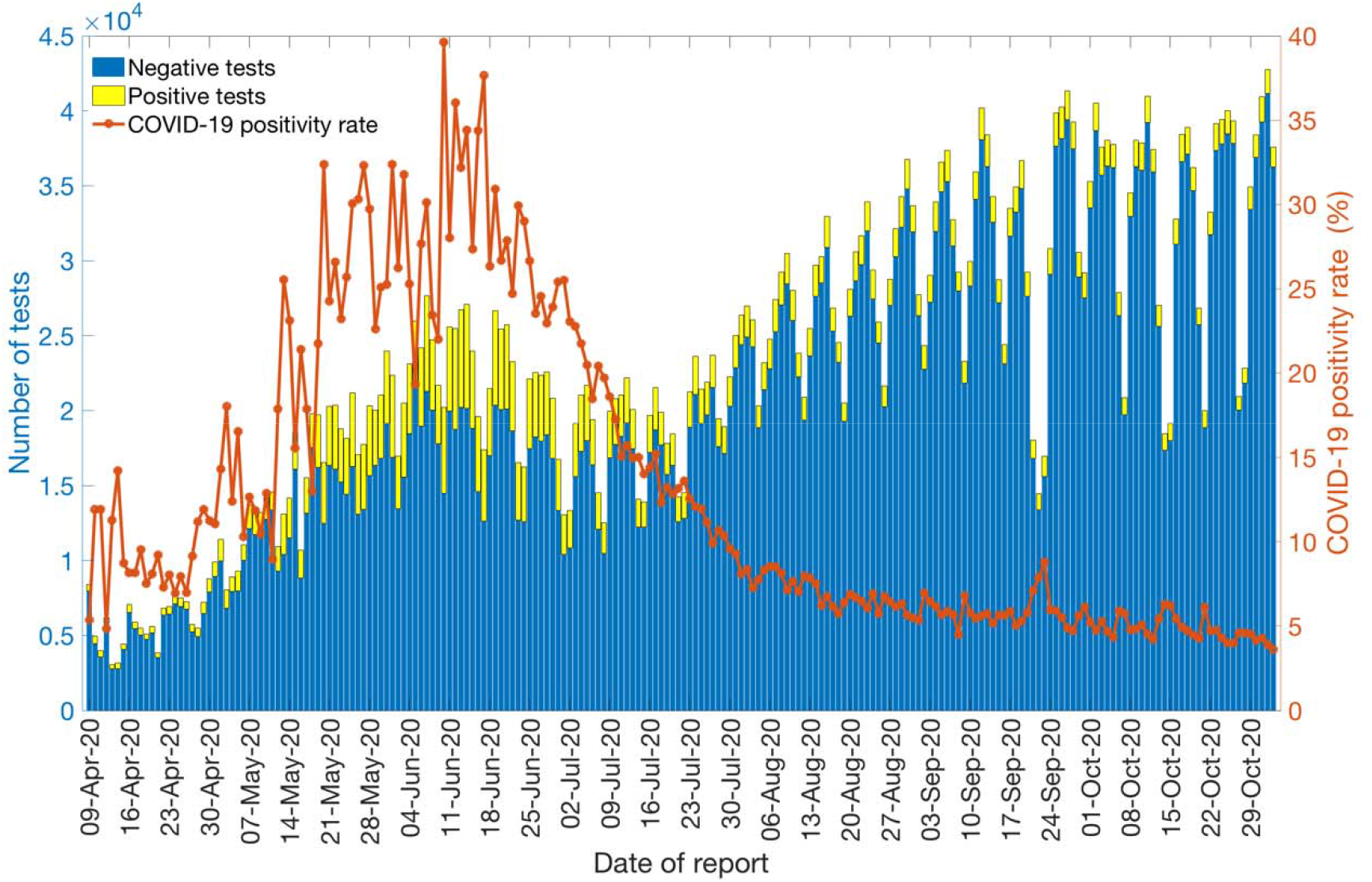
Laboratory results for the COVID-19 tests conducted in Chile as of November 2^nd^, 2020. The blue color represents the negative test results, and the yellow color represents the positive test results. The solid orange line represents the positivity rate of COVID-19 in Chile.

## 4. Discussion

The estimates of the early transmission potential in Chile for the first 27 days of the epidemic indicate sustained local transmission in the country with the estimate of reproduction number *R* at ∼1.8 (95% CI: 1.6, 1.9) which is also in accordance with the estimate of the reproduction number obtained from the Cori method (R∼2.2 95% CrI (2.14, 2.28)). The estimates of *R* from our analysis agree with the estimates of *R* retrieved from studies conducted in the surrounding Latin American countries including Peru and Brazil [71, 72]. Other countries including Korea, South Africa and Iran also exhibit similar estimates of *R* that lie in the range of 1.5-7.1 [73-80]. In contrast, some other countries including Singapore and Australia have reported much lower estimates of *R* (*R* <1) that can be correlated with the implementation of early strict social distancing interventions in these countries [81, 82].

The initial deceleration of the growth parameter in Chile indicates a sub-exponential growth pattern (*p*∼0.8), consistent with sub-exponential growth patterns of COVID-19 that have been observed in Singapore (*p*∼0.7), Korea (*p*∼0.76) and other Chinese provinces excluding Hubei (*p*∼0.67) [78, 81, 83]. In contrast, studies conducted in Peru, a Latin American country, and Iran have reported a nearly exponential growth pattern of the COVID-19 whereas an exponential growth pattern has been reported in China [72, 75, 83].

Although the initial transmission stage of COVID-19 in Chile has been attributed to multiple case importations, Chile quickly implemented control measures against the COVID-19 epidemic, including border closures on March 18^th^, 2020, to prevent further case importations. The 20-day ahead forecast of our GGM model calibrated to 28 days suggest that the social distancing measures, including closure of schools, universities and day cares, have helped slow down the early virus spread in the country by reducing population mobility (Table 1, Figure 1, Figure 6) [84]. The commixture of interventions, including localized lockdowns, night-time curfew, school closures, and the ban on social gatherings in Chile, can probably be attributed to preventing the disease trend from growing exponentially during the early growth phase, as has occurred elsewhere [3, 4]. However, the significant increase in case incidence observed in mid-May can probably be attributed to the relaxation of social distancing measures and reopening of society in late April, in the context of the “Safe Return” plan [31]. As the virus reached the lower-income neighborhoods in Chile, the pandemic quickly exploded [23, 38, 39, 85]. While the COVID-19 case incidence exhibited a relative stabilization in case trajectory for April 2020 (with an average of ∼443 cases per day), highlighting the positive effects of early quarantine and lockdowns, the reopening of society and early economic reactivation in late April 2020 probably resulted in the surge of cases resulting in an acceleration of the epidemic with estimates of *R* higher than 1.0. The total lockdown comprised of stay-at-home orders imposed in Greater Santiago (which accounted for about 77% of cases in the country) on May 15^th^ showed an effect in slowing the virus’s transmission. Similar lockdowns were imposed in Antofagasta on May 5^th^ and in Valparaíso on June 12^th^, though these regions together represent only ∼10% of cases in Chile [31]. The deceleration of growth parameter, *p*, has been estimated at ∼0.51 (95% CI: 0.47, 0.56) after the reimposition of lockdowns and social distancing measures in May, consistent with a linear incidence growth trend, indicating deceleration of the epidemic.

Moreover, we estimated a reproduction number, *R*, of ∼0.87 (95% CI: 0.84, 0.89) in early July, indicating a decline in transmission of the virus consistent with the stay-at-home orders. This reproduction number corresponds to the instantaneous reproduction number estimated during the course of the epidemic utilizing the Cori method, which also indicates a decrease in disease transmission with *R*∼0.8 as of early July. The instantaneous reproduction number has fluctuated around ∼1 since early August with the most recent estimate of reproduction number, *R*∼0.9 as of November 2^nd^, 2020. The 20-day ahead forecast calibrating data to the GLM model (from May 9^th^-July 7^th^, 2020) has reasonably indicated a declining trend in case incidence. The forecast results also imply that approximately ∼45,160 cases (95% CI: 27,934-67,600) could be observed in Chile from July 8^th^-July 27^th^, 2020. The actual case count by for this time period, as reported by the Chilean government indicated 46,798 cases, closely approximating our mean GLM forecast, falling within the 95% PI. Therefore, based on the most recent estimates of *R* (Figure 5), it can be implied that maintaining social distancing measures, limiting social gatherings, and reducing population mobility have served as essential ways to containing the spread of the virus [86, 87].

Though the number of reported cases in Latin America remains low compared to the United States, official data for many Latin American countries are incomplete. However, Chile has tested a higher percentage of its residents than any other Latin American nation, lending confidence to its reliability [88]. Chile’s testing capabilities have been greatly expanded during the pandemic, in part from a coordinated effort lead by the Ministry of Science to include testing from public and private laboratories. For instance, the average number of COVID-19 tests performed in Chile per day per thousand people is 1.52 compared to the neighboring South American country, Peru (∼0.2 tests per thousand people) [89]. The average positivity rate for the whole span of the epidemic in Chile is estimated at ∼12.98%. However, the average monthly positivity rate of COVID-19 in Chile is estimated at ∼5.90% and ∼4.88% for September and October, respectively, compared to ∼20.09% in May 2020. The high positivity rate at the beginning of the epidemic indicates that the government failed to cast a wide enough net to test the masses early in the pandemic, and there were probably many more active cases than those detected by epidemiological surveillance, underestimating the epidemic growth curve [90-92]. The most recent lower testing rates indicate that Chile is testing a comparatively larger segment of the population. This positivity rate for Chile is also consistent with the positivity rate obtained from India, the United States, Canada, and Germany that exhibit moderately high positivity rates (4-8%) for COVID-19, indicating overall limited testing in these countries [89, 93]. In comparison, some countries like Mexico and the Czech Republic exhibit very high positivity rates (30-51%) [89]. Other countries like Denmark, New Zealand, Australia, Singapore, and South Korea have reached very low positivity rates (0-3%) by testing the masses with South Korea’s large testing capacity combined with a strategy that tracks infected people via cell phones [88, 89, 94]. Moreover, studies suggest there is asymptomatic transmission of SARS-CoV-2 [66, 95, 96], which means we could have underestimated our estimates based on the daily incidence’s growth trend from symptomatic cases [97-99]. On the other hand, preliminary results of a study have shown the relative transmission of asymptomatic cases in Santiago to be almost ∼3% [100]. While our study highlights the effectiveness of broad-scale social distancing and control interventions in Chile, it also underscores the need for persistent isolation and social distancing measures to stomp all active disease transmission chains in Chile. In the absence of pharmacological interventions and considering the advent of second waves in Asia and Europe, non-pharmacological interventions such as the ones implemented in Chile are the available options for countries to address the pandemic before large segments of the population are immunized with effective and safe vaccines. In this scenario, real-time metrics that characterize the transmission dynamics and control are crucial to face the future challenges that the pandemic will impose.

This study has some limitations. First, our study analyzes cases by the dates of reporting while it is ideal to analyze the cases by the dates of onset or after adjusting for reporting delays. On the other hand, a substantial fraction of the COVID-19 infections exhibits very mild or no symptoms at all, which may not be reflected by data [101, 102]. Moreover, the data are not stratified by local and imported cases; therefore, we assumed that all cases contribute equally to the transmission dynamics of COVID-19. Finally, the extent of selective underreporting, and its impact on these results is difficult to assess.

## 5. Conclusions

In this study, we estimate the transmission potential of SARS-CoV-2 in Chile. Our current findings point to sustained transmission of SARS-CoV-2 in the early phase of the outbreak, with our estimate of the reproduction number at *R*∼1.8. The COVID-19 epidemic in Chile followed an early sub-exponential growth trend (*p* ∼0.8) which has transformed into an almost linear growth trend (*p* ∼0.5) as of July 7^th^, 2020. The most recent estimate of reproduction number, *R*, is ∼0.9 as of November 2^nd^, 2020, indicating a decline in the virus transmission in Chile. The implementation of lockdowns and apt social distancing interventions have indeed slowed the spread of the virus. However, the number of new COVID-19 cases continue to accumulate, underscoring the need for persistent social distancing and active contact tracing efforts to maintain the epidemic under control.

## Data Availability

All data are publicly available.

## Author Contributions

Conceptualization, G.C. and A.T.; methodology, G.C, A.T.; validation, G.C.; formal analysis, A.T., G.C.; investigation, A.T.; resources, G.C., A.T.; data curation, A.T.; writing— original draft preparation, A.T., G.C.; writing, review and editing, A.T., G.C., E.U, C.C, K.V., R.R, R.L, G.C.; visualization, A.T., G.C.; supervision, G.C.; project administration, G.C.; funding acquisition, G.C. All authors have read and agreed to the published version of the manuscript.

## Availability of data and materials

The datasets used and analyzed during the current study are available from Base de Datos COVID-19 repository, http://www.minciencia.gob.cl/covid19.

